# Genotype-First Assessment of Presentation and Penetrance of Neurofibromatosis Type 1, Autosomal Dominant Polycystic Kidney Disease, and Marfan Syndrome Within the *All of Us* Research Program Cohort

**DOI:** 10.1101/2025.02.26.25322940

**Authors:** Stephanie A Felker, Bruce R. Korf, Gregory S. Barsh

**Author notes:** CORRESPONDENCE Gregory S. Barsh.

## Abstract

**Purpose:** Phenotype-based ascertainment of probands in studies of Mendelian disorders may exclude individuals with mild phenotypes or that lack health care access. We explore this premise in *All of Us* Research Program participants with pathogenic variation causal for three Mendelian conditions: autosomal dominant polycystic kidney disease (ADPKD), Marfan syndrome, and neurofibromatosis type 1 (NF1).

**Methods:** We identified *All of Us* Research Program participants with putatively pathogenic variation in *NF1*, *FBN1*, *PKD1*, and *PKD2*. Concept terms were extracted from electronic health records to assess participant diagnosis and phenotype. Variant annotation and participant surveys were evaluated to identify biological and social factors differentiating diagnosed and undiagnosed individuals.

**Results:** Large proportions of individuals with pathogenic variation in *NF1*, *FBN1*, or *PKD1/PKD2* lack the associated diagnosis of NF1 (47%), Marfan syndrome (58%), or ADPKD (51%), respectively. Pathogenic variants in diagnosed individuals have greater inferred deleteriousness for NF1 and ADPKD, and undiagnosed individuals had less severe phenotypes compared to diagnosed individuals for all three conditions.

**Conclusion:** A genotype-first ascertainment of individuals in genomic research allows for a more comprehensive assessment of Mendelian disease and removes biases that confound our understanding of the penetrance and presentation of these conditions.

## INTRODUCTION

Our understanding of Mendelian conditions has historically been based on probands identified on the basis of their clinical and/or laboratory findings, accompanied by examination and investigation of their relatives. This phenotype-based approach has provided considerable insight into presentation, natural history, penetrance, and expressivity, but may be incomplete for several reasons. Individuals who do not have a clinically observable phenotype or cannot access comprehensive medical care are categorically excluded, and studies may be biased towards individuals with a more prominent disease phenotype. Much of the clinical genetic literature comes from academic medical centers in which some groups are underrepresented due to race, gender, ethnicity, or social determinants of health. In addition, biological and social contributors may be intertwined. As population-based genome sequencing becomes increasingly available, a genotype-first approach offers potential new insight into our understanding of mendelian conditions, and may reveal confounders to disease diagnosis, including non-biological factors such as race, gender, ethnicity, and access to health insurance.

The *All of Us* Research Program is a NIH program that enrolls individuals, performs genome sequencing, and links electronic health record (EHR), survey, and demographic data to enable research [1]. As of the *All of Us* Curated Data Repository (CDR) version 7 (participant cut-off July 1, 2022), this data includes 245,388 individuals with short read whole genome sequencing (srWGS), 182,459 of which have clinical diagnoses in their linked EHR. The program emphasizes enrollment of individuals historically underrepresented in biomedical research and includes extensive survey data detailing social determinants of health, income, education, and healthcare access and utilization. We used this resource to carry out a genotype-first assessment of three Mendelian conditions: autosomal dominant polycystic kidney disease (ADPKD), Marfan Syndrome, and neurofibromatosis type 1 (NF1). These conditions were chosen because they are relatively prevalent (1/1000 – 1/10,000), thought to be highly penetrant based on family studies, and >90% of affected individuals for each condition have mutations in only one (*NF1*, *FBN1*) or two (*PKD1*, *PKD2*) genes [2, 3, 4].

Our work reveals that, for each of the three conditions as ascertained by genotype, penetrance is less than expected and the extent of variable expressivity is greater than expected. We identify biologic (e.g. inferred deleteriousness) and social (e.g. race, access to health care) and factors associated with lack of a diagnosis despite the presence of a pathogenic Mendelian variant. These results have important implications for our understanding of the natural history of, and genetic counseling for, these conditions when identified by genome sequencing rather than phenotype.

## MATERIALS AND METHODS

### All of Us Research Program Cohort and Data Acquisition

This investigation was conducted using the EHR, survey, demographic, and srWGS data available through the *All of Us* Research Program Researcher Workbench. Data from 182,459 individuals with both srWGS and at least one medical diagnosis within *All of Us* CDR v7 were analyzed. A joint SNP/indel callset of all srWGS data was obtained via a Hail VariantDataset (VDS) file accessible through the *All of Us* CDR, and EHR data was acquired on the *All of Us* Researcher Workbench using a custom Concept Set created that included any clinical finding according to Systemized Nomenclature of Medicine – Clinical Terms (SNOMED CT) [5]. These codes were collated for each of the 182,459 individuals as described above, and the number of SNOMED CT for each individual was determined, with significance between cohorts determined by a Wilcoxon rank-sum test conducted in the R stats package [6]. Cohorts of participants with ADPKD, Marfan syndrome, and NF1 were determined by the presence of their associated SNOMED CT or daughter concepts using the *All of Us* Researcher Workbench Cohort Builder. Concept terms included 2 SNOMED CT for ADPKD: 765330003 (Autosomal dominant polycystic kidney disease), 253880009 (Autosomal dominant polycystic kidney disease in childhood); 1 SNOMED CT for Marfan syndrome: 19346006 (Marfan syndrome); and 3 SNOMED CT for NF1: 92824003 (Neurofibromatosis type 1), 403816002 (Multiple cafe-au-lait macules due to neurofibromatosis), and 403817006 (Multiple neurofibromas in neurofibromatosis).

### Variant Curation within PKD1, PKD2, FBN1, and NF1

*All of Us* participants consenting to srWGS data provide whole blood samples at *All of Us* recruitment centers, which are sequenced and genotyped at *All of Us* Genome Centers. SrWGS data is aligned to the GrCh38 genome and is subjected to rigorous QC with regard to single-sample, joint call set, and batch effects as previously detailed [7]. All variants in *PKD1*, *PKD2*, *FBN1*, and *NF1* (MANE transcript coordinates chr16:2088708-2135898, chr4:88007635-8807777, chr15:48408313-48645709, and chr17:31094977-31377675, respectively) were acquired from the *All of Us* CDR v7 srWGS VDS and annotated via Ensembl’s Variant Effect Predictor (VEP) REST API version 15.7 [8, 9]. Variants with either a Pathogenic or Likely Pathogenic assignation in ClinVar [10], “High Confidence” score using LoFTEE [11], or a VEP “HIGH” biological impact score were considered putatively pathogenic. Variants with an allele balance below 0.30 or above 0.70 were excluded to select only confidentially heterozygous germline variants. The minimum read depth of all resulting variants positions was 14, therefore no additional depth filtering was conducted. Additionally, as a conservative measure, post-hoc curation of variants was conducted to discount any variant that was found exclusively in over 5 undiagnosed individuals, which included *PKD1* variants NM_001009944.3:c.12391G>T, NM_001009944.3:c.10337del, and NM_001009944.3:c.9829C>T. Hereafter, individuals with putatively pathogenic variants will be referred to as “variant-positive.” The Combined Annotation-Dependent Depletion (CADD) score [12] for each variant was obtained in the annotation process described above, and the median putatively pathogenic variant CADD score of diagnosed and undiagnosed cohorts for each condition was compared using a Wilcoxon rank sum test.

### Interrogation of Common Variants in Undiagnosed Individuals

We recognize that assigning pathogenicity via previous ClinVar submissions and computational predictions of biological consequence may errantly incorporate benign variants due to overestimated computational predictions or empirical variant misinterpretations. To that end, we identified common variants that were exclusively found in subsets of undiagnosed individuals (Supplemental Table 1, Entries 1-6). Additionally, there are variants that are seen in both undiagnosed and diagnosed individuals (Supplemental Table 1, Entries 7-10), suggesting that these variants are pathogenic, yet these individuals remain undiagnosed.

### EHR Analysis

The number of SNOMED CT codes present in the EHR of each participant was collected from the *All of Us* Researcher Workbench. We compared median SNOMED CT count of “variant-positive” diagnosed and undiagnosed cohorts for each condition as well as the *All of Us* Research Program as a whole. The median age of “variant-positive” diagnosed and undiagnosed individuals was extracted and compared to the All of Us Research Program median age. Additionally, as above-average height is a clinical feature of Marfan syndrome, the sex-normalized z-scored height of *FBN1* variant-positive diagnosed and undiagnosed individuals was extracted and compared to the All of Us Research Program median z-scored height.

Condition-specific phenotype risk scores (PheRS) were calculated for each of the 182,459 individuals [13] for ADPKD, NF1, and Marfan syndrome. PheRS is a weighted-aggregated approach where the clinical features for each condition are scored (w_p_) based on their prominence in the *All of Us* Research Program:

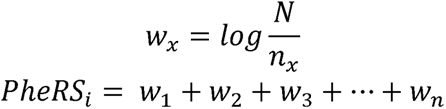

Where *w_x_* is the score for each clinical feature of the condition, *N* is the total number of individuals in the cohort examined (182,459), *n_x_* is the number of individuals with the clinical feature within the cohort, and PheRS_i_ is the sum of the *w_x_* for clinical features present in an individual’s EHR, which is the individual’s PheRS score for the condition. Clinical features for each condition were accumulated from the Online Mendelian Inheritance in Man (OMIM [14]) clinical synopsis for each condition (Phenotype MIM numbers: NF1: 162200, PKD1: 173900, PKD2: 613095, FBN1: 154700), and only clinical features with a corresponding SNOMED CT present in the *All of Us* Research Program were used. For each clinical feature, the SNOMED CT for the feature and all daughter concepts determined by https://browser.ihtsdotools.org/ [15], using the 07-31-2021 International Version SNOMED CT dictionary in accordance with *All of Us* Research Program v7 CDR (Supplementary Table 2). Significant difference was determined using a Wilcoxon Rank Sum Test.

### Demographics Analysis

Upon enrollment in All of Us, participants are given the opportunity to complete surveys assessing demographic and lifestyle data. Self-reported values for race, gender, date of birth, and ethnicity were obtained from the pre-built “Demographics” Concept Set, health insurance status and type were obtained through the pre-built “Healthcare Access and Utilization” dataset (specifically question concept ID “1585386” and “43528428”), and height data was acquired by creating a concept set including the “Body height” concept ID (3036277). Significant difference in “variant-positive” diagnosed and undiagnosed cohorts in categorical values (race, ethnicity, gender) were determined via Fisher’s Exact test, and significant differences in continuous variables (age and height) were determined via a Wilcoxon rank sum test.

## RESULTS

### Diagnostic Overview and EHR Results

Within the *All of Us* Research Program CDR v7, there are 182,459 individuals with srWGS and at least one EHR-derived medical diagnosis. We obtained a srWGS Hail VariantDataset SNP/indel call set for these individuals through the Controlled Tier Dataset within the *All of Us* Researcher Workbench. For EHR data, we created a custom Concept Set that included any clinical finding according to Systemized Nomenclature of Medicine – Clinical Terms (SNOMED CT) [5]. Diagnosis of ADPKD, Marfan syndrome, and NF1 were determined by the presence of their associated SNOMED CT or daughter concepts.

For genome variation within *PKD1*, *PKD2*, *FBN1*, or *NF1*, we combined computational predictions (Variant Effect Predictor, LoFTEE) with expert curation (ClinVar) to produce a comprehensive set of putatively pathogenic variants in *All of Us* participants. Additional quality control metrics were developed and applied as described in Materials and Methods to exclude false positives and somatic mosaicism.

We identified a total of 471 *All of Us* participants with putatively pathogenic variants in *PKD1* (133), *PKD2* (39), *FBN1* (120), and *NF1* (179), results that are consistent with the prevalence of these conditions: ADPKD (1 in 1000), Marfan syndrome (1 in 5,000 - 10,000), and NF1 (1 in 2,000 - 4,000). Although each of these conditions is thought to be highly penetrant, 51%, 58%, and 47% of individuals with pathogenic variation in *PKD1*/*PKD2*, *FBN1*, and *NF1*, respectively, did not have a SNOMED CT coding for the condition or daughter concept of the condition in their *All of Us* EHR (Figure 1B).

**Figure 1:**
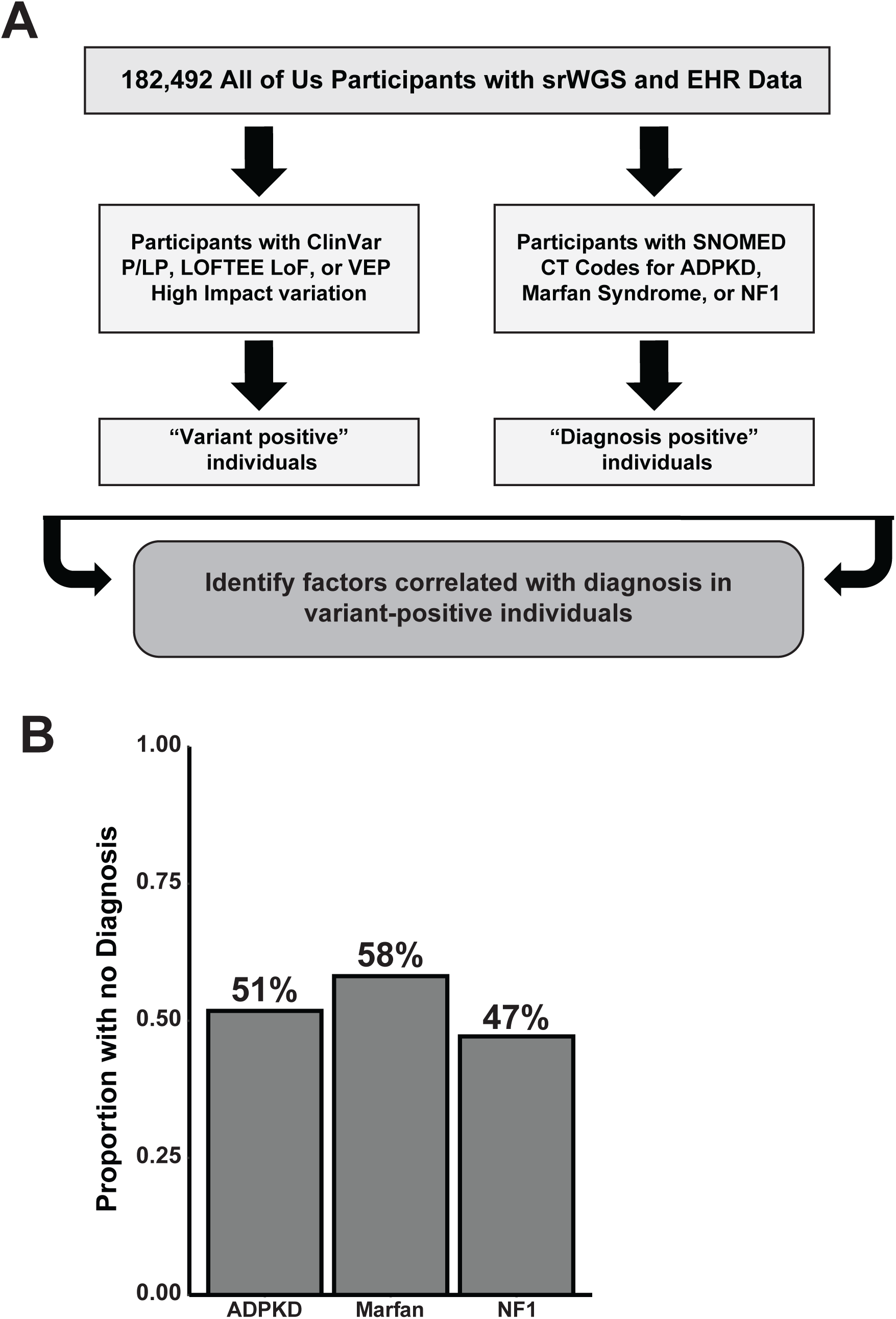
Large proportions of individuals have pathogenic variation without diagnosis of three autosomal dominant Mendelian conditions. A: Schematic of methodology implemented to identify individuals and analyze with diagnosis of genetic disease and/or pathogenic variation within the *All of Us* Research Program. B: Proportion of individuals with pathogenic variation in genes associated with ADPKD (n = 137), Marfan Syndrome (n = 43), and NF1 (n=123) without a diagnosis of the relevant condition.

We considered several potential explanations for the surprising reduction of apparent penetrance, including deficiencies in the EHR, variable expressivity, and differences in SDoH. In what follows, we refer to the groups of individuals with putatively pathogenic variation but no corresponding EHR diagnosis as “undiagnosed”. Potential deficiencies in the EHR were evaluated by examining and comparing the number of SNOMED CT codes between different groups (Figure 2A). For PKD and Marfan syndrome, the median number of SNOMED CT codes for undiagnosed individuals is similar to and not significantly different from the average *All of Us* participant (42 for PKD, 37 for Marfan syndrome, and 37 for the average *All of Us* participant). For NF1, undiagnosed individuals have a smaller and marginally significant (26 vs. 37, p = 0.0355, Wilcoxon rank sum test) number of SNOMED CT codes than the average *All of Us* participant. For all three conditions, diagnosed individuals have more SNOMED CT codes than undiagnosed individuals (Figure 2A). Taken together, these results indicate that EHR differences may contribute to apparent reduced penetrance for NF1 and point to variable expressivity as a contributing factor for apparent reduced penetrance in all three conditions.

**Figure 2:**
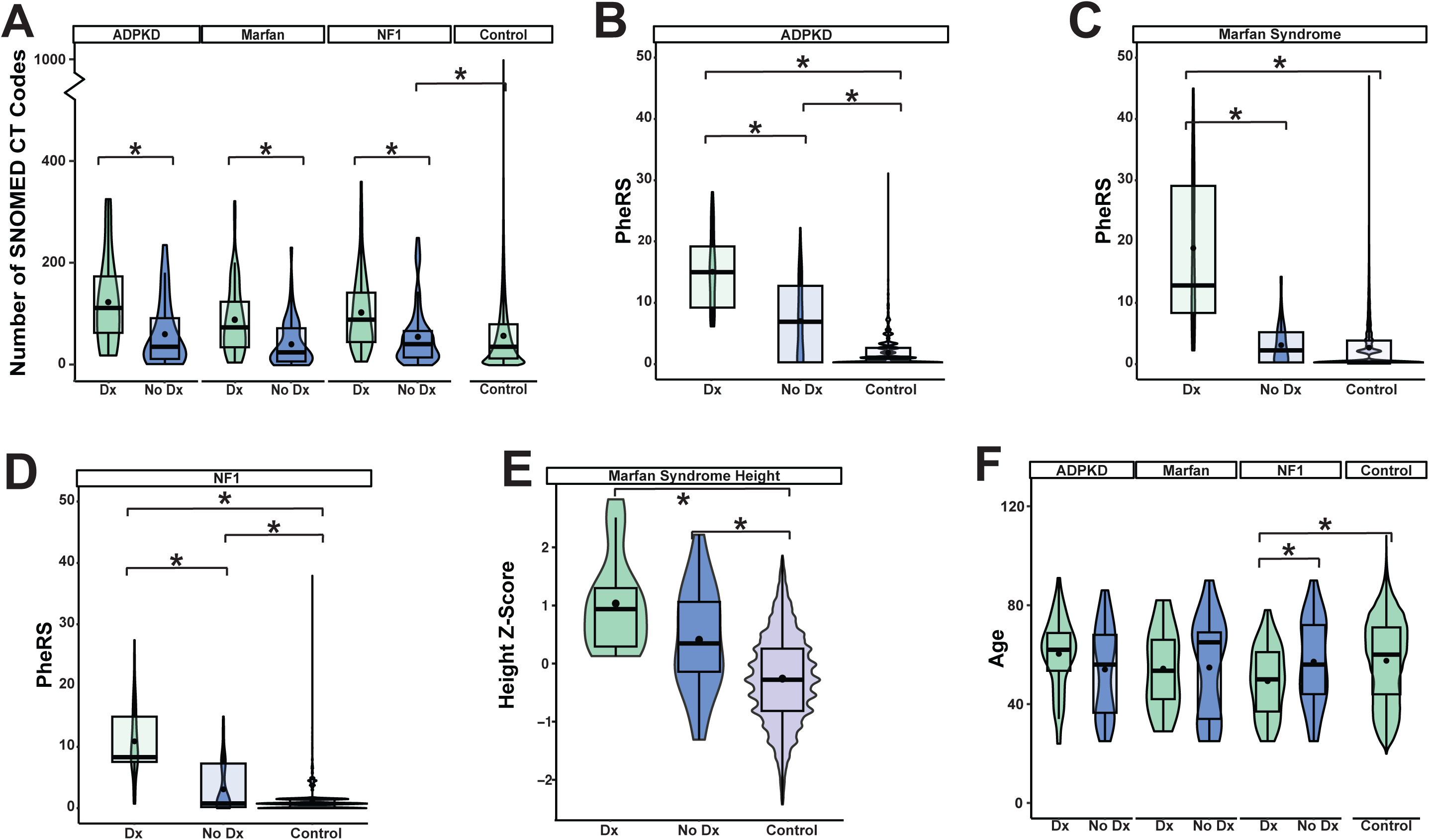
EHR and Clinical comparison of variant-positive individuals with and without a diagnosis of Mendelian conditions. A: Number of SNOMED CT codes in *All of Us* Research Program EHR for diagnosed (Dx) and undiagnosed (No Dx) individuals (ADPKD: diagnosed vs. undiagnosed p = 7.996e-06, undiagnosed vs. control p = 0.7819; Marfan syndrome: diagnosed vs. undiagnosed p = 0.008098, undiagnosed vs. control p = 0.8665; NF1: diagnosed vs. undiagnosed p = 3.212e-06, undiagnosed vs. control, p = 0.0355). PheRS scores for ADPKD (B: diagnosed vs. undiagnosed p = 7.367e-10, undiagnosed vs. control p = 1.079e-11), Marfan syndrome (C: diagnosed vs. undiagnosed p = 1.128e-06, undiagnosed vs. control p = 0.241), and NF1 (D: diagnosed vs. undiagnosed p = 1.187e-13, undiagnosed vs. control p = 0.002211). “Control” individuals are remaining of the *All of Us* Research Program participants, without a pathogenic variant or a diagnosis of associated condition. E: Height of individuals diagnosed and undiagnosed with Marfan Syndrome (diagnosed vs. undiagnosed p = 0.02925, undiagnosed vs. control p = 0.0003048). All participants’ height was normalized by gender via z-score. F: Age of diagnosed and undiagnosed individuals for each condition (ADPKD: diagnosed vs. undiagnosed p = 0.06189, undiagnosed vs. control p = 0.09417; Marfan syndrome: diagnosed vs. undiagnosed p = 0.1865; undiagnosed vs. control p = 0.9992; NF1 diagnosed vs. undiagnosed p = 0.0154, undiagnosed vs. control p = 0.693). * = p-values ≤ 0.05 determined via Wilcoxon rank sum test.

### Variable expressivity: PheRS Analysis and Height

Phenotype risk scores (PheRS) are a quantitative metric that represents the extent to which an individual’s phenotype or set of phenotypes matches a well-characterized Mendelian condition. We calculated condition-specific PheRS scores as described in Materials and Methods and compared their distributions between diagnosed and undiagnosed individuals for PKD, Marfan syndrome, and NF1 (Figures 2B-D). For all three conditions, undiagnosed individuals had median PheRS scores that were significantly less than diagnosed individuals (ADPKD 14.9 vs. 6.71, p = 7.367e-10; Marfan syndrome 12.7 vs. 1.99, p = 1.128e-06; NF1 8.31 vs. 0.77, p = 1.187e-13; Wilcoxon rank sum test). Also, for all three conditions, undiagnosed individuals had PheRS scores that were higher than the average *All of Us* participant although the difference for Marfan syndrome was not significant (ADPKD 6.7 vs. 0.77, p = 1.079e-11; Marfan syndrome 1.99 vs. 0, p = 0.241; *NF1* 0.77 vs. 0.73 p = 0.002211, Wilcoxon rank sum test). We also note that, for all three conditions, the PheRS distributions for both the diagnosed and undiagnosed groups are broad and overlapping (Figure 2B-D). In other words, individuals with an EHR diagnosis exhibit a considerable range of phenotypic expressivity, and among those without an EHR diagnosis there are some individuals who have the same or greater PheRS score as the median for individuals with an EHR diagnosis. For Marfan syndrome, we also examined height as a potential proxy for the extent of variable expressivity. As depicted in Figure 2E, the median height of undiagnosed individuals is intermediate between that of diagnosed individuals and the average *All of Us* participant (undiagnosed vs. diagnosed p = 0.02925; diagnosed vs. control p = 1.447e-08; undiagnosed vs. control p = 0.0003048).

### Relationship of Age to Penetrance

The clinical features of NF1 and Marfan syndrome are usually recognized in childhood and adolescence, respectively, but ADPKD often does not come to clinical attention until later in life. For ADPKD (Figure 2F), comparison of age distributions between diagnosed and undiagnosed groups showed a small effect of median age that was marginally significant (62 vs. 56 p = 0.06056). There was no effect of age on apparent penetrance of Marfan syndrome, but for NF1, the median age of the diagnosed group was lower than the undiagnosed group (50 vs. 56, p = 0.01161) and also lower than the average *All of Us* participant (50 vs. 60, p = 3.805e-05).

### Variant Deleteriousness is Correlated with Apparent Penetrance

Lower PheRS scores in undiagnosed individuals could reflect differences in the mutation spectrum if the variants in the diagnosed groups are more deleterious than those in the undiagnosed groups. In addition, differences in SDoH and/or demography may contribute to lower PheRS scores if those individuals have less access to medical care. To examine the former possibility, we compared the CADD scores of pathogenic variants found in diagnosed versus undiagnosed individuals (Figure 3A). We found that variants in diagnosed individuals had higher CADD scores than the variants in the respective undiagnosed cohorts, with a significant difference in ADPKD and NF1 cohorts (ADPKD p = 0.01695; Marfan Syndrome p = 0.9636; NF1 p = 0.003705). Additionally, we found that diagnosed individuals have greater proportions of variants with high biological consequences (frameshift, splice effects, start loss/stop gained) than individuals without a diagnosis in ADPKD (p = 0.03148), Marfan Syndrome (p = 0.03448) and NF1 (p = 0.004498) (Supplemental Figure 2).

**Figure 3.**
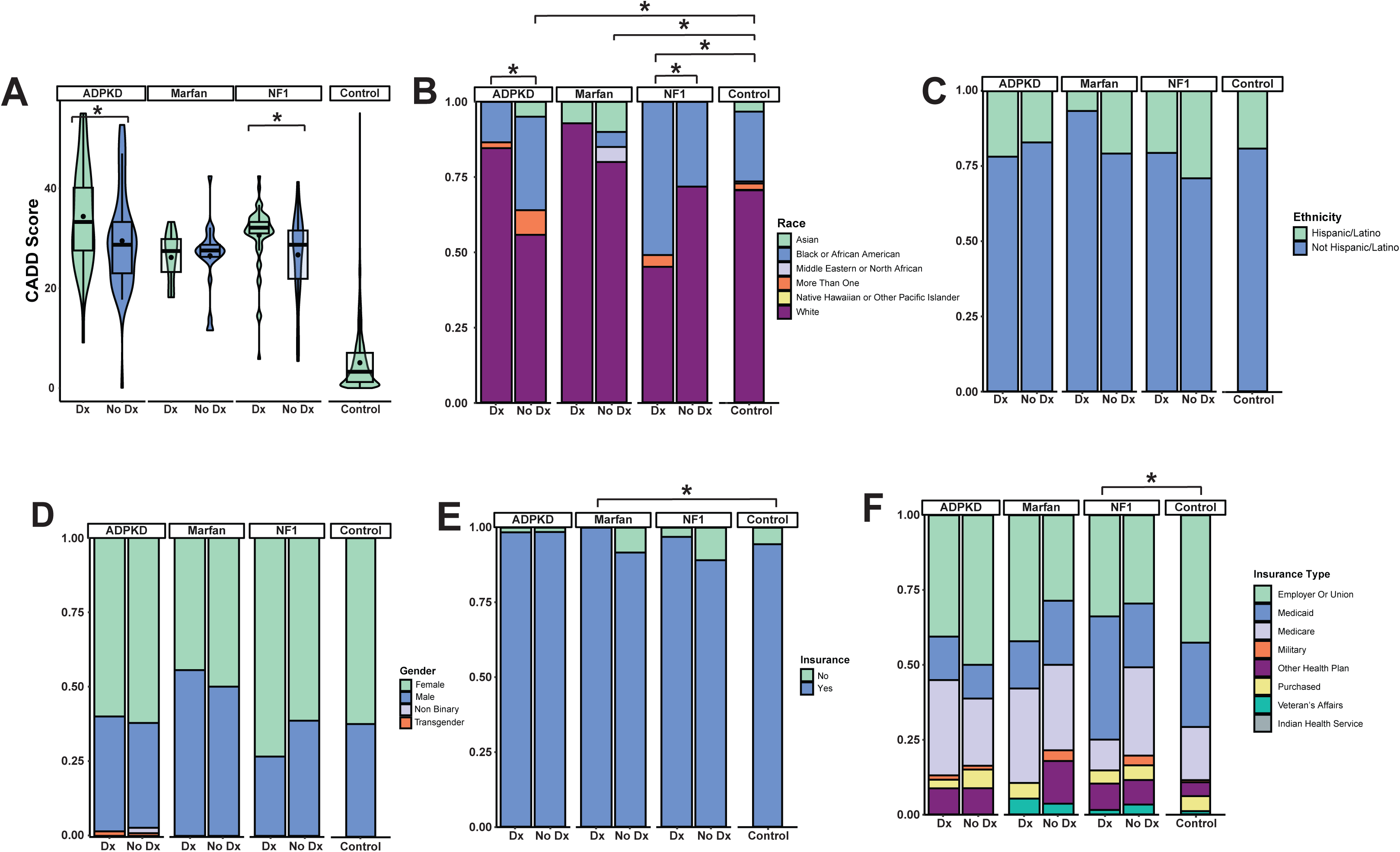
Social and biological demographics for variant-positive diagnosed vs. undiagnosed individuals. A: CADD score of pathogenic variants found in diagnosed and undiagnosed individuals * p-values ≤ 0.05 via Wilcoxon rank sum test (ADPKD: diagnosed vs. undiagnosed p = 0.0003825; Marfan syndrome: diagnosed vs. undiagnosed p = 0.3238; NF1 diagnosed vs. undiagnosed p = 0.004699). B-F Proportions of self-reported race (B, ADPKD: diagnosed vs. undiagnosed p = 0.05953, diagnosed vs. control p = 0.1639, undiagnosed vs. control p = 0.06347; Marfan syndrome diagnosed vs. undiagnosed p = 0.8181, diagnosed vs. control p = 0.9185, undiagnosed vs. control p = 0.2249; NF1: diagnosed vs. undiagnosed p = 0.04505, diagnosed vs. control p = 0.0004998, undiagnosed vs. control p = 0.6802), ethnicity (C, ADPKD: diagnosed vs. undiagnosed p = 0.6999, diagnosed vs. control p = 1, undiagnosed vs. control p = 0.3916; Marfan syndrome: diagnosed vs. undiagnosed p = 0.1801, diagnosed vs. control p = 0.3371, undiagnosed vs. control p = 0.3685; NF1: diagnosed vs. undiagnosed p = 0.514, diagnosed vs. control p = 0.629, undiagnosed vs. control p = 0.1607), gender (D, ADPKD: diagnosed vs. undiagnosed p = 1, diagnosed vs. control p = 0.6961, undiagnosed vs. control p = 0.6224; Marfan syndrome: diagnosed vs. undiagnosed p = 0.1756, diagnosed vs. control p = 0.3187, undiagnosed vs. control p = 0.4695; NF1: diagnosed vs. undiagnosed p = 0.1171, diagnosed vs. control p = 0.08901, undiagnosed vs. control p = 0.5773), health insurance status (E, ADPKD: diagnosed vs. undiagnosed p = 0.4255, diagnosed vs. control p = 0.2611, undiagnosed vs. control p = 1; Marfan syndrome: diagnosed vs. undiagnosed p = 1, diagnosed vs. control p = 0.6214, undiagnosed vs. control p = 0.6239; NF1: diagnosed vs. undiagnosed p = 0.07567, diagnosed vs. control p = 0.4377, undiagnosed vs. control p = 0.03656), and health insurance type (F, ADPKD: diagnosed vs. undiagnosed p = 0.7964, diagnosed vs. control p = 0.1559, undiagnosed vs. control p = 0.1119; Marfan syndrome: diagnosed vs. undiagnosed p = 0.239, diagnosed vs. control p = 0.6807, undiagnosed vs. control p = 0.5082; NF1 diagnosed vs. undiagnosed p = 0.02247, diagnosed vs. control p = 0.006997, undiagnosed vs. control p = 0.7316) in diagnosed and undiagnosed individuals, compared to the whole of the *All of Us* Research Program (”Control”). * p-values ≤ 0.05 determined via Fisher’s Exact test.

### Demographic and Social Differences between Diagnosed and Undiagnosed Individuals

We examined whether apparent reduced penetrance for ADPKD, Marfan syndrome, and NF1 is correlated with race (Figure 3B), ethnicity (Figure 3C), gender (Figure 3D), and health insurance status/type (Figure 3E and 3F). We found significant differences between racial distributions in diagnosed and undiagnosed individuals with pathogenic *PKD1* and *PKD2* variants (p = 0.00474) and undiagnosed individuals and the entire *All of Us* cohort (p = 0.01199). There was also a significant difference in the racial distribution between individuals with pathogenic *FBN1* variation lacking a diagnosis and the control cohort (p = 0.02849). We additionally found that proportions of race in NF1-diagnosed individuals vs. undiagnosed individuals were significantly different (p = 0.0224) and NF1-diagnosed individuals were significantly different from the *All of Us* cohort (p = 0.0004998).

With regards to ethnicity, across all three conditions, the diagnosed individuals had a greater proportion of non-Hispanic/Latino individuals, but these differences did not reach significance for any condition (ADPKD, diagnosed vs. undiagnosed p = 0.5189, diagnosed vs. control p = 0.6331, undiagnosed vs. control 0.7625; Marfan Syndrome, diagnosed vs. undiagnosed p = 0.3758, diagnosed vs. control p = 0.3307, undiagnosed vs. control 0.7967; NF1, diagnosed vs. undiagnosed p = 0.3916, diagnosed vs. control p = 0.7494, undiagnosed vs. control 0.0841).

A potential relationship between apparent penetrance and race could reflect biologic and or social differences. The majority of *All of Us* participants have health insurance; however, we observed that for Marfan syndrome and NF1, there was a higher proportion of undiagnosed compared to diagnosed individuals that lacked health insurance (Figures 3E-F). Those differences are not statistically significant, although for Marfan syndrome, the proportion of individuals with health insurance is higher in the diagnosed group compared to the average *All of Us* participant (p = 0.0003241). At a more granular level, the diagnosed group for NF1 had a significantly different distribution of health insurance type compared to both undiagnosed (p = 0.02822) and control (0.01049) individuals.

## DISCUSSION

As genome sequencing becomes increasingly affordable, the way in which we recognize, diagnose, care for, and counsel patients with Mendelian conditions will evolve. As described here, our findings indicate that penetrance of 3 Mendelian conditions when ascertained by genotype is substantially less than when ascertained by phenotype.

In this work we refer to *All of Us* participants with putatively pathogenic variation in *PKD1*, *PKD2*, *FBN1*, or *NF1* but who do not have EHR evidence of the corresponding conditions as “undiagnosed”. Because the proportion of undiagnosed individuals for all three conditions was much larger than anticipated, we considered two major types of confounding explanation.

First, one or more of the variants in the undiagnosed group may have been benign because of a curation mistake (ClinVar) or a false positive computational prediction. For these reasons, we excluded any variant that was only reported once in ClinVar or that was observed in 5 or more undiagnosed individuals. As depicted in Supplemental Figure 1, there was considerable overlap between the two platforms we used for computational prediction but less overlap with ClinVar variants. Overall, putatively pathogenic variants from ClinVar that lacked computational support accounted for a greater proportion of Marfan syndrome individuals than for ADPKD or NF1 individuals, both diagnosed and undiagnosed. While this observation could reflect more ClinVar curation errors in Marfan syndrome, a more likely explanation is that unique structural requirements of the fibrillin protein can be disrupted by missense variants that are not classified as pathogenic by LOFTEE or VEP (Supplemental Figure 2).

A second potential source of confounding is that that some individuals in the undiagnosed group may have had inaccurate and/or incomplete records in the *All of Us* EHR. However, we note that the median number of SNOMED CT codes for undiagnosed individuals across all three conditions was 32 (range 1-250), similar to the average *All of Us* participant (median 37, range 1-988). And, while diagnosed individuals had, overall, a higher median number of SNOMED CT codes than undiagnosed individuals, the ranges between the diagnosed and undiagnosed groups exhibited considerable overlap. Finally, undiagnosed individuals, exhibited phenotypic characteristics as assessed by PheRS scores or height for Marfan syndrome, that were significantly greater than the average *All of Us* participant. Taken together, these considerations indicate that, overall, lack of an EHR diagnosis in individuals with pathogenic variation in *PKD1*, *PKD2*, *FBN1*, or *NF1* is due to a lower extent of phenotypic expressivity.

From a patient’s perspective, our results show that there are many individuals with pathogenic variation in ADPKD, Marfan syndrome, or NF1 that may benefit from additional medical care. From a practitioner’s perspective, our results call into question how health care providers interpret, explain, and provide genetic counseling for individuals with apparently pathogenic variation but who have mild and/or few symptoms. Many of these questions will be addressed with additional studies to, e.g., determine the extent to which undiagnosed individuals with pathogenic *FBN1* variants develop serious sequelae that would benefit from surveillance and intervention. As population-based genome sequencing becomes increasingly applied, the utility of the information will depend on repositories like *All of Us* that facilitate integration with an EHR that is both comprehensive and longitudinal.

## DATA AVAILABILITY

### Supplementary Data and Supplementary Methods

All data was accessed and analyzed via *All of Us* Researcher Workbench using Jupyter Notebooks in the “Examining the Effect of SDoH on the Diagnosis of Mendelian Conditions” searchable in *All of Us* Researcher Workbench.

## Data Availability

All data produced are available online through the All of Us Researcher Workbench

https://workbench.researchallofus.org

## ACKNOWLEDGEMENTS

We thank all the participants of the *All of Us* Research Program. SAF is supported by NHGRI 5T32HG008961.

## AUTHOR CONTRIBUTIONS

Conceptualization: S.A.F., B.R.K., G.S.B.; Data curation: S.A.F.; Formal analysis: S.A.F.; Funding acquisition: S.A.F.; Investigation: S.A.F.; Methodology: S.A.F.; Project administration: S.A.F., G.S.B., Resources: B.R.K., G.S.B.; Software: S.A.F.; Supervision: G.S.B.; Visualization: S.A.F.; Writing-original draft: S.A.F., G.S.B.; Writing-review & editing: S.A.F., B.R.K., G.S.B.

## ETHICS DECLARATION

We exclusively used data that is accessible through the *All of Us* Researcher Workbench. As stated on the *All of Us* website, all participants have consented to be involved in the *All of Us* Research Program, including data from electronic health records (EHRs), surveys, and physical measurements. All data was generated and shared in accordance with the *All of Us* Data Statistics Dissemination Policies.

---

**Supplementary Figure 1.**
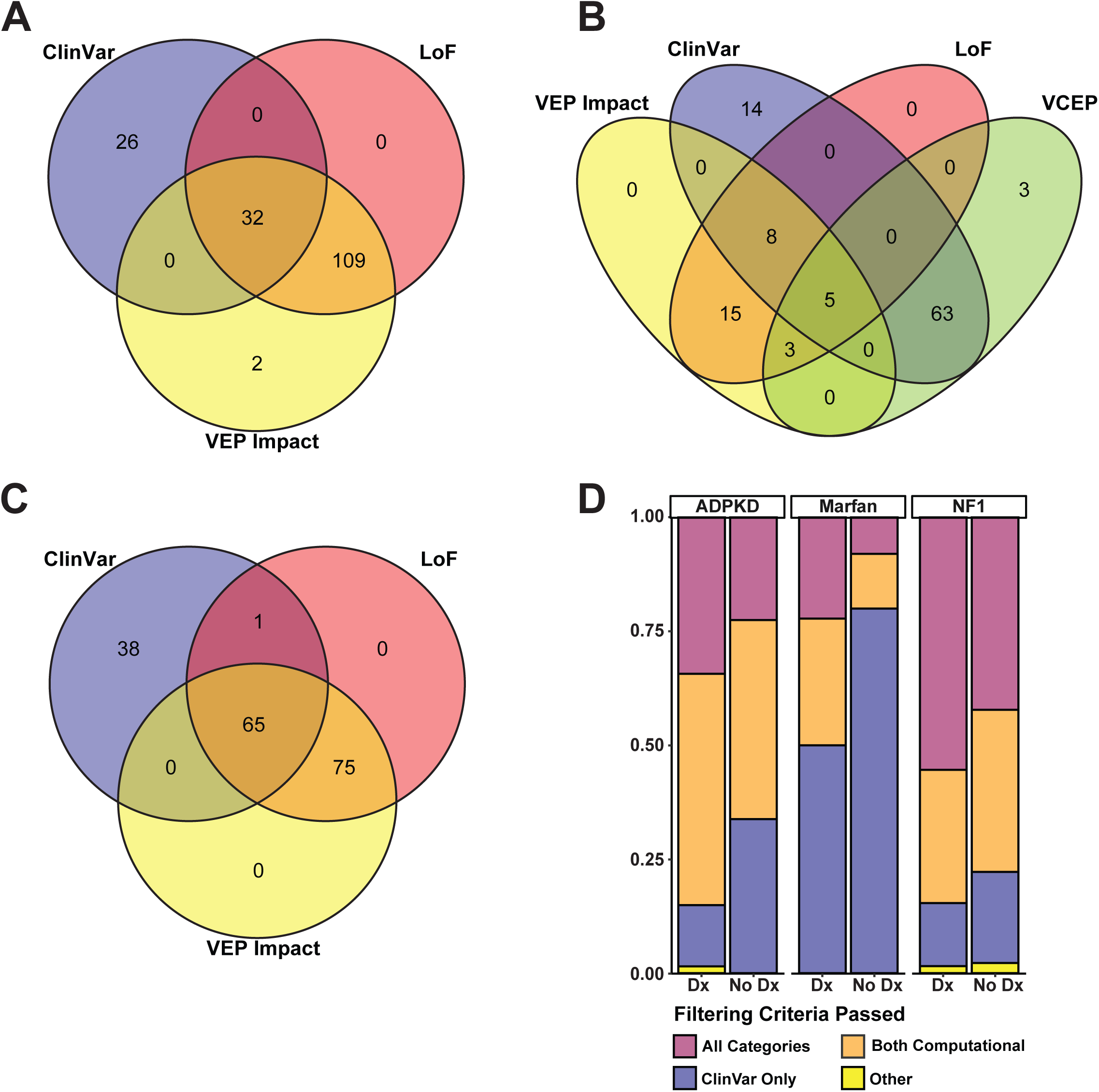
Overview of Pathogenicity Criteria in Variants in PKD1, PKD2, FBN1, and NF1. Variants were determined to be pathogenic if they were designated Pathogenic or Likely Pathognic in ClinVar (ClinVar; total = 208 variants), assigned “High Confidence” LoF using LoFTEE annotation (LoF; total = 308 variants), or had a “HIGH” VEP Impact score (VEP Impact; total = 311 variants). A: PKD1 and PKD2 (169 total variants), B: FBN1 variants (111 total variants), C: NF1 Variants (179 total variants). D: Proportions of individuals with pathogenic variants passing all, computational, or ClinVar only annotation filtering criteria.

**Supplemental Figure 2.**
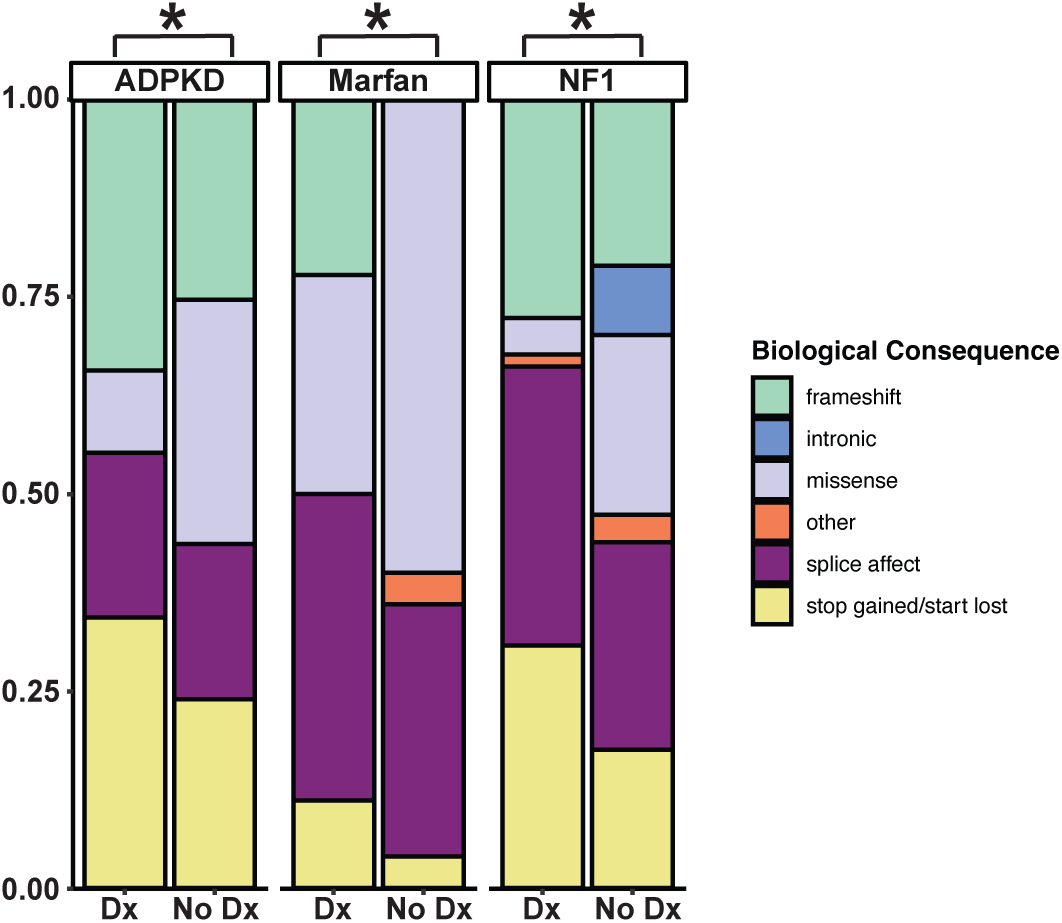
Biological consequence of pathogenic variation found in diagnosed vs. undiagnosed individuals. Biological consequences as predicted rom VEP annotations. * = p-values ≤ 0.05 determined via Fisher’s Exact test (ADPKD diagnosed vs. undiagnosed p = 0.02349; Marfan syndrome diagnosed vs. undiagnosed p = 0.03048; NF1 diagnosed vs. undiagnosed p = 0.001999).

**Table 1.**
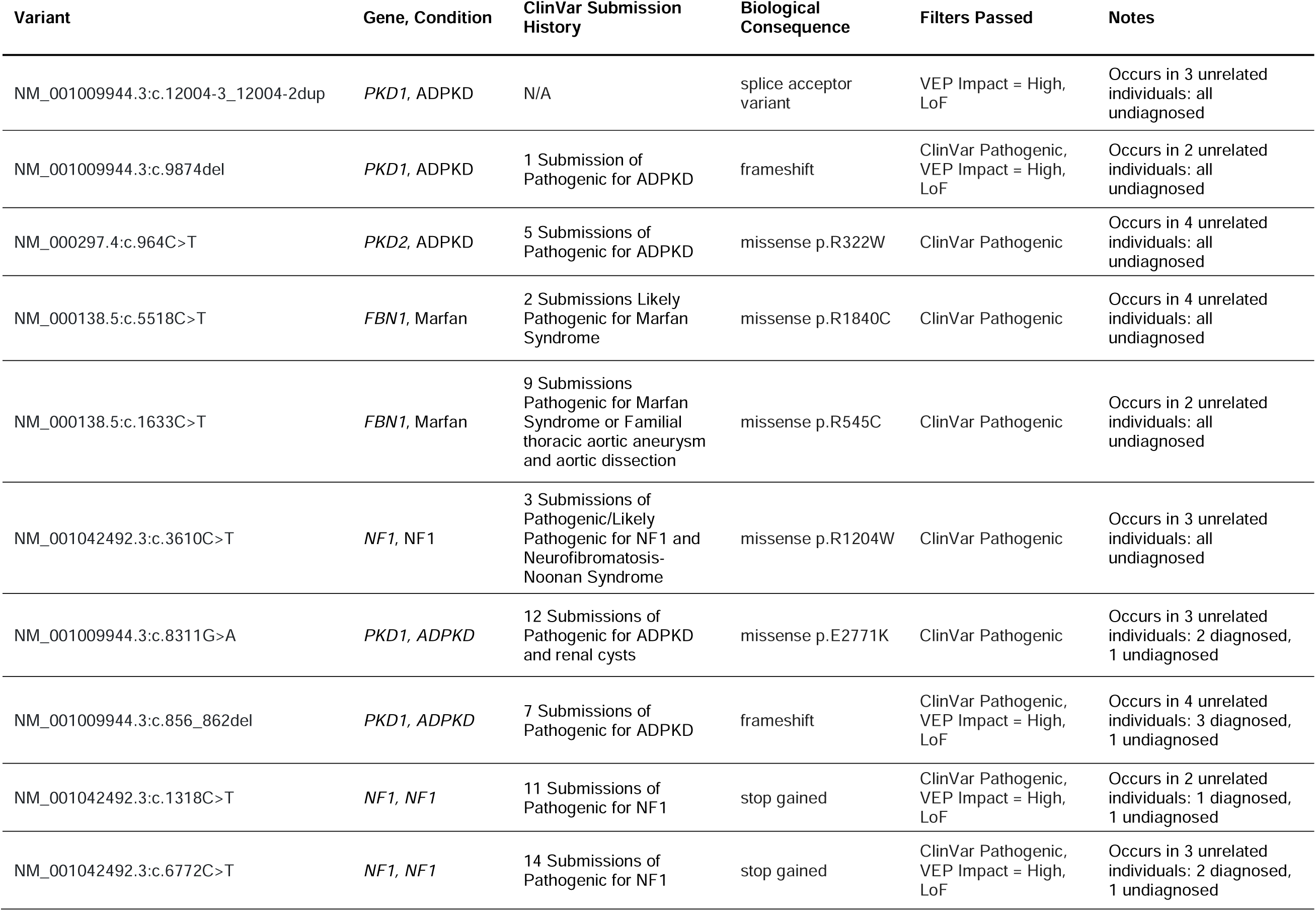
Common Variants in Undiagnosed Individuals.

